# Reconstructing the first COVID-19 pandemic wave with minimal data in the UK

**DOI:** 10.1101/2023.03.17.23287140

**Authors:** Siyu Chen, Jennifer A Flegg, Katrina A Lythgoe, Lisa J White

## Abstract

Accurate measurement of exposure to SARS-CoV-2 in the population is crucial for understanding the dynamics of disease transmission and evaluating the impacts of interventions. However, it is particularly challenging to achieve this in the early phase of a pandemic because of the sparsity of epidemiological data. In our previous publication[1], we developed an early pandemic diagnostic tool that can link minimum datasets: seroprevalence, mortality and infection testing data to estimate the true exposure in different regions of England and found levels of SARS-CoV-2 population exposure are considerably higher than suggested by seroprevalence surveys. Here, we re-examined and evaluated the model in the context of reconstructing the first COVID-19 epidemic wave in England from three perspectives: validation from ONS Coronavirus Infection Survey, relationship between model performance and data abundance and time-varying case detection rate. We found that our model can recover the first but unobserved epidemic wave of COVID-19 in England from March 2020 to June 2020 as long as two or three serological measurements are given as model inputs additionally, with the second wave during winter of 2020 validated by the estimates from ONS Coronavirus Infection Survey. Moreover, the model estimated that by the end of October in 2020 the UK government’s official COVID-9 online dashboard reported COVID-19 cases only accounted for 9.1% (95%CrI (8.7%,9.8%)) of cumulative exposure, dramatically varying across two epidemic waves in England in 2020 (4.3% (95%CrI (4.1%, 4.6%)) vs 43.7% (95%CrI (40.7%, 47.3%))).

## Introduction

The COVID-19 pandemic has inflicted devastating effects on global populations and economies [2, 3] and now still affecting countries in many different ways. Reviewing the challenges posted by the COVID-19 pandemic and evaluating previous responses is vital important for future pandemic preparedness [4-7]. Accurate estimation of exposure remains crucial for understanding the dynamics of disease transmission and assessing the impacts of interventions along different stages of pandemic. However, this is particularly challenging in the early phase since most of the characteristics of the pathogen are unknown and at the same time epidemiological data are sparse.

Confirmed COVID-19 cases was typically the first type of data to be collected and reported mostly due to the syndrome surveillance systems [8, 9]. However, it usually underestimates the true exposure in the population because of the limited capacity of diagnoses, the unsolid definition of cases, testing criteria and etc. Large-scale viral infection survey in the community can help to solve the testing issue. For example, the UK Office for National Statistics (ONS) conducted a national wide COVID-19 viral testing survey, namely Covid Infection Survey (CIS) [10] that successfully tracked the trajectories of COVID-19 infections in the community of UK since April of 2020. Because of its representative sampling across households in the general population this study is recognised to have a strong power to capture asymptomatic infections which might be missed out by symptomatic testing scheme in the early pandemic and can provide reliable estimates of prevalence over time [11]. However, this study started collecting samples from April of 2020 and then reporting the estimates of daily incidence from May of 2020 while the first death due to COVID-19 disease in the UK was documented in February 2020 [12]. This implies that the transmission of COVID-19 in the community began earlier than the survey, and the survey might not be able to recover the early epidemic curve.

Serologic studies that measure how many people have antibodies against the virus are a promising tool for pinning down the stage of the pandemic because of its ability of capturing past infections regardless of clinical symptoms [13]. If the antibody elicited by the virus lasts for lifetime, representative sampling in a population followed by the antibody testing will provide robust estimates of exposure. However, cohort studies following individuals over time after they’ve had a known COVID-19 infection were able to determine that antibodies are only measurable up to 6–9 months [14-16], on average, varying across testing assay [17] and antigen types [18]. The immediate implication is that serological studies will inevitably under-estimate the number of people exposed, since some will have a lower antibody count when the study is conducted and test negative. Linking multiple publicly available datasets, we proposed a method that have been published previously [1] to estimate the true level of exposure after considering the antibody decay. Here we further examined and evaluated the model in the context of reconstructing the first COVID-19 pandemic from three perspectives: validation from ONS Infection Survey, relationship between model performance and data abundance and time-varying case detection rate.

## Result

### Reconstruction of the early epidemic

In our previous paper, we presented a simple model to link together three key metrics for evaluating the progress of an epidemic, applied to the context of SARS-CoV-2 in England: antibody seropositivity, infection incidence and number of deaths. We use these three metrics to estimate the antibody seroreversion rate and region-specific infection fatality ratios. In doing so, the cumulative number of infections in England are estimated, showing that cross-sectional seroprevalence data underestimate the true extent of the SARS-CoV-2 epidemic in England in the early pandemic. Estimates for the IgG (spike) seroreversion rate and IFR are broadly consistent with other studies, which supports the validity of these findings.

The model was set up based on the important observation about the COVID-19 infection timeline that seroconversion in individuals who survive occurs at approximately the same time as death for those who do not. Therefore, a simple ordinary differential equation (ODE) was formulated to model the rate of change in the number of seropositive individuals in different regions of England which will increase as new infections were generated that was calculated by the daily number of deaths dividing by infection fatality ratio and will decrease as antibody decay. The model predicted seropositive population were fitted to observed seroprevalence using a Bayesian observation model.

### Validation from ONS Infection Survey

Comparing the incidence of SARS-CoV-2 in England estimated by our model with those inferred by ONS Coronavirus Infection Survey (Figure 1), we found that our model could reveal the first but unobserved epidemic wave of COVID-19 in England from March 2020 to June 2020 additionally, with the second wave validated by the estimates from ONS Infection Survey. Further, we found our model results were highly consistent with those using SEIRS type compartmental models with time-varying force of infection [19, 20].

**Figure 1.**
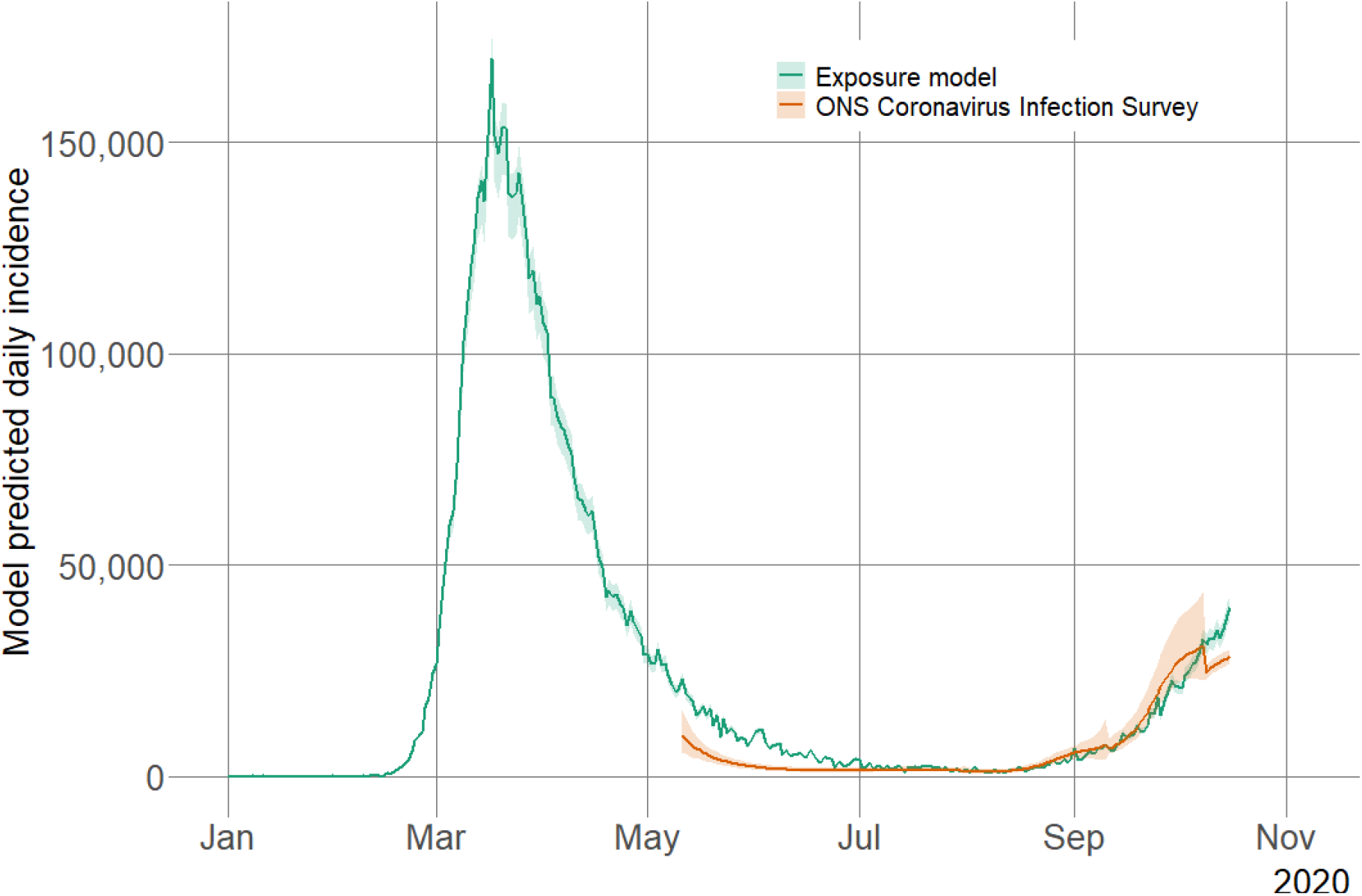
Comparison of model predicted daily incidence of SARS-CoV-2 in England. The green lines show the predictions of median daily incidence by our model [1] based on Equation (1) and (2) in the Materials and Methods section. The orange lines show the predictions of median daily incidence from ONS Coronavirus Infection Survey while the orange shaded areas correspond to the 95% CrI.

### Relationship between model performance and data abundance

We then examined the relationship between model performance and data abundance - how estimates of exposure from our model change with more serological data points being added into the fitting procedure one by one over time (Figure 2). We found a highly robust pattern of exposure across different regions of England was estimated in general.

**Figure 2.**
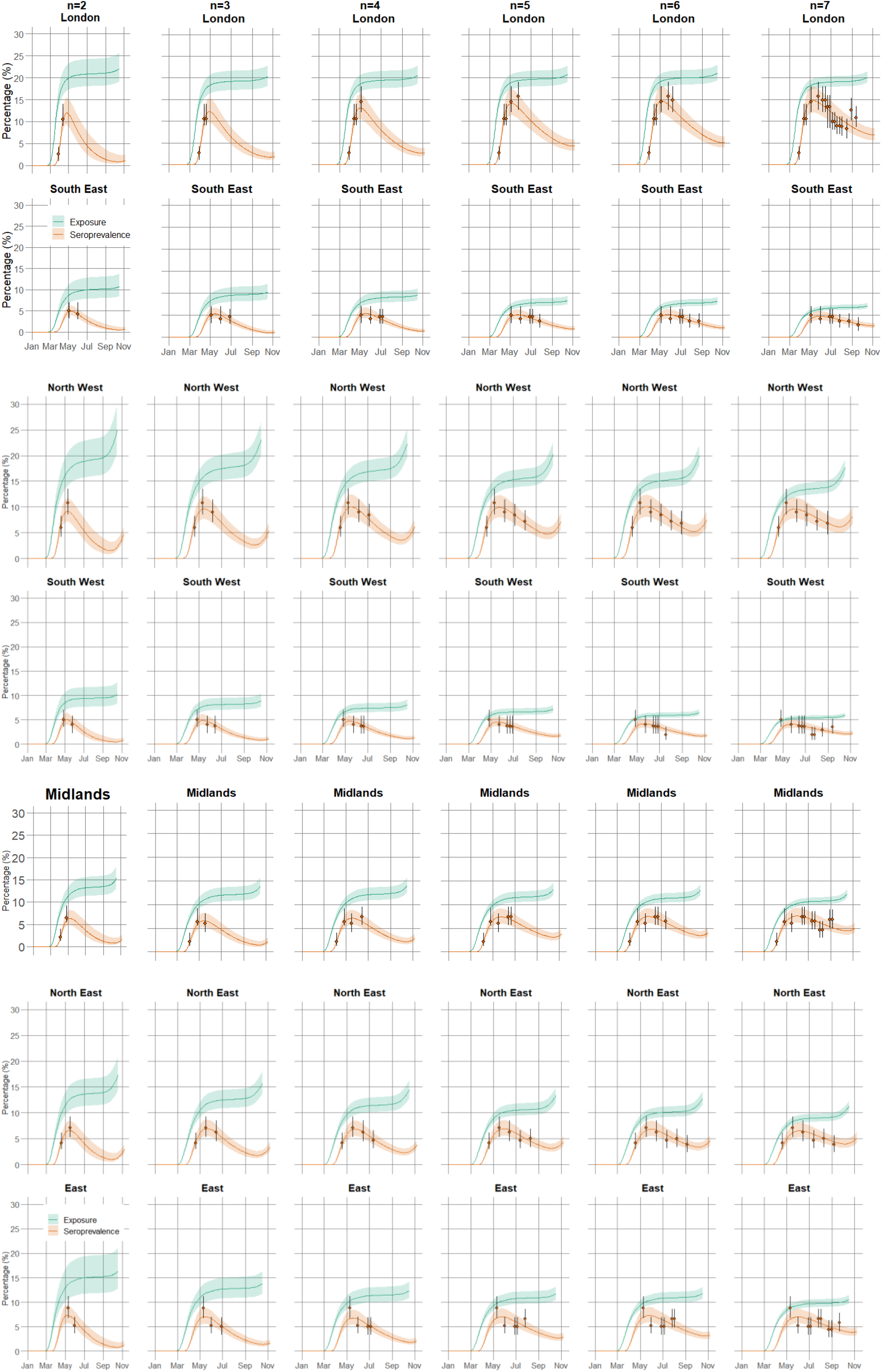
Comparison of estimates of exposure in seven regions of England as more serological measurements are given as model inputs (left to right). The green and orange lines show the model predictions of median exposure and seroprevalence, respectively, while the shaded areas correspond to the 95% CrI.

Specifically, the model could only start estimating the interested quantities: exposure and two parameters (infection fatality ratio and antibody decaying rate) when at least two serological measurements from April to June 2020 in each region were given as inputs. However, these estimates were already highly consistent with those when more serological measurements were added although the credible bands were wider. The wide credible bands suggested a bigger uncertainty around the estimates when little information was available. When three serological measurements in each of region were included the estimates of exposure level became largely consistent at the results when all serological measurements were used. This might be attributed to the timing of these third serological measurements since then the seroprevalence in most regions started decreasing. With more and more serological measurements being added, the credible bands of estimates of exposure gradually narrowed.

### Time-varying case detection rate

While comparing the reported cases with the incidence estimated by our model (Figure 3), we found the UK government’s official COVID-9 online dashboard (https://coronavirus.data.gov.uk) reported COVID-19 cases in England only accounted for 9.1% (95%CrI (8.7%,9.8%)) of cumulative exposure by the end of October 2020. Further, the relative size of two infection waves in England in 2020 estimated by our model, Spring wave from February to June and Autumn wave from September to November, were reversed compared those reported by the confirmed cases. The case detection rate relative to the total exposure was also dramatically different in these two-epidemic waves. If separating the two waves from the first of August 2020, we found during January 2020 to August 2020 the case detection rate was only 4.3% (95%CrI (4.1%, 4.6%)) which increased to 43.7% (95%CrI (40.7%, 47.3%)) during August 2020 to October 2020, highlighting the dominate effect of testing effort in shaping the case curve in the early stage of a pandemic. The testing issue, e.g. the limited capacity of tests and symptom-based testing strategy posted a big challenge for understanding the early pandemic. Viral surveys in the general population can solve the sampling issue, but still have the problem of not sampling early on. Serological data even from some convenient samples, e.g., blood donors can help to pin down the progress of the pandemic when antibody decay is teased out.

**Figure 3.**
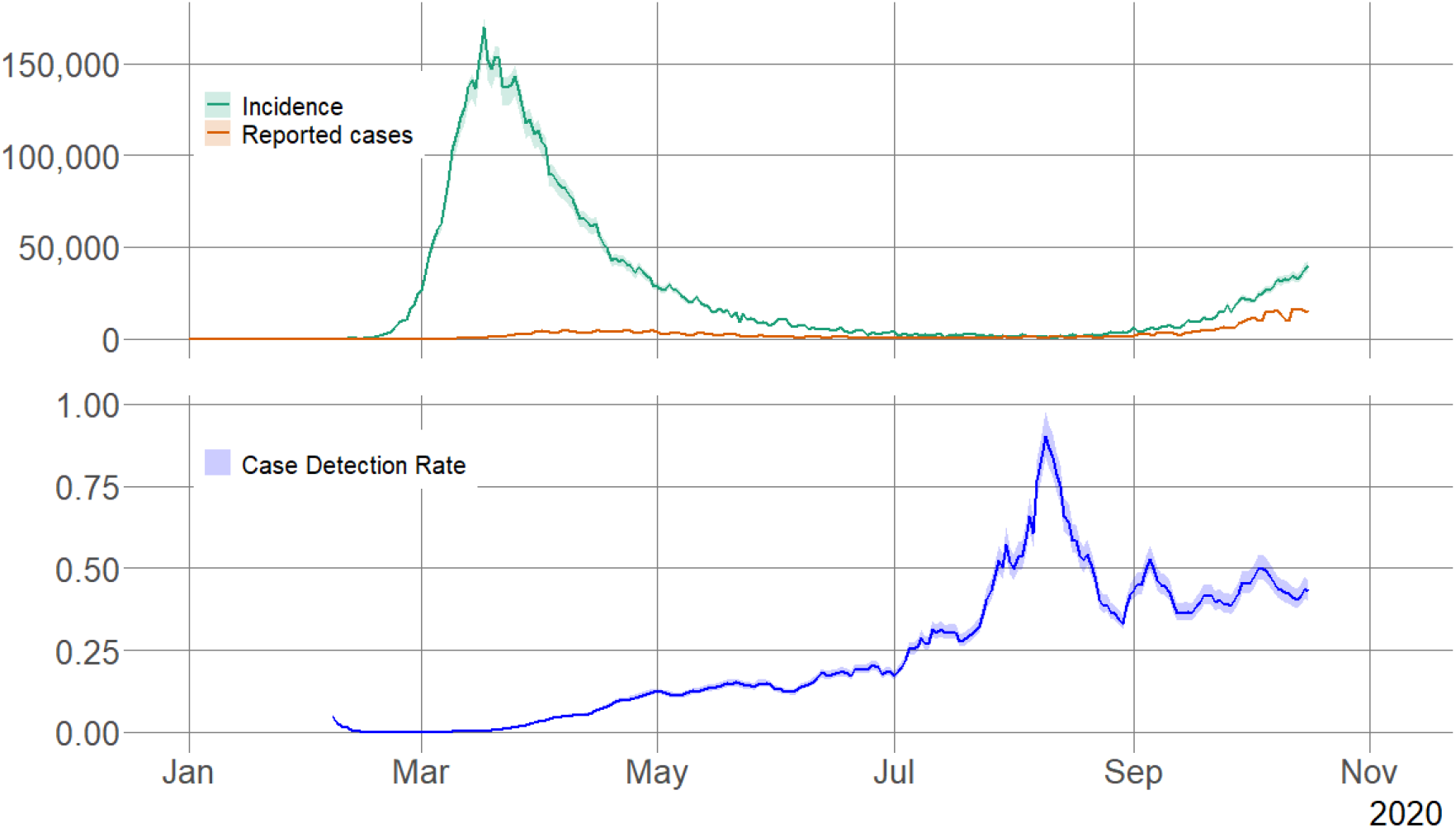
Comparison between estimates of daily incidence with reported cases of SARS-CoV-2 in England and case detection rate. Here, all serological measurements were used in the model fitting. In the top figure, the green lines show the predictions of median daily incidence by our model based on Equation (1) and (2) in the Materials and methods section while the shaded areas correspond to the 95% CrI. The red lines show the reported confirmed cases in England downloaded from GUV.UK dashboard. In the bottom figure, the blue lines show the estimates of median case reporting rate in England based on Equation (3) and (4) while the shared areas correspond to the 95% CrI.

## Discussion

Accurate reconstruction of exposure time series is necessary to assess how policies influenced transmission over time, in particular when reporting is lagged, and multiple interventions may have been undertaken in succession. For example, [21] made use of the comparison of exposure between general population and pregnant women in New York City to conclude the effectiveness of shielding during pregnancy. Moreover, the prior exposure level in the population can be used to inform future intervention design, e.g., vaccination prioritisation. For example, in the early stage of the COVID-19 vaccination campaign, when dose supply and administrative capacity were initially limited worldwide, a modelling study [22] explored how uncertainty about previous exposure levels and about a vaccine’s characteristics affects the prioritization strategies for reducing deaths and transmission. This model showed use of individual-level serological tests to redirect doses to seronegative individuals improved the marginal impact of each dose while potentially reducing existing inequities in COVID-19 impact.

Here, we evaluated a simple dynamic model that we published previously and demonstrated its ability of reconstructing the first epidemic wave before large-scale survey sampling by providing robust estimates of exposure over time. One key element of the model was fitting model to serologic data that was generated from healthy adult blood donors supplied by the NHS Blood and Transplant (NHS BT collection) serum samples using the Euroimmun anti-spike IgG assay and reported in the Weekly national Influenza and COVID-19 surveillance report. This suggests that convenient samples, for example here serum samples from blood donors have the promising power to provide primary information of epidemic progress in a short timeframe especially during the emergency of a new outbreak from a novel pathogen.

Because of the rigorous sampling design and robust estimation power ONS Covid Infection Survey can almost be seen as a golden standard for estimating community prevalence. Our model does not take any results or estimates from the survey as inputs, so the comparison exercise that we conducted here between estimates of exposure from our model with ONS Covid Infection Survey provides a real-world validation. Moreover, we showed the modelling approach is a valuable early pandemic diagnostic tool and can clearly recover the first epidemic wave that the survey was unable to capture because of late starting time. Using the inferred daily incidence, we explicitly demonstrated the variation of case detection rates over two epidemic waves in England in 2020. It provides quantitative information for studying the association between the capacity, behaviour, strategy of testing with the epidemic evolution and further supported the argument that confirmed cases largely underestimate the extent of disease transmission.

Moreover, the simple structure of the presented model avoids unnecessary complexity and structure-based uncertainty in a full dynamic model where compartmental models simulating the disease spread in different groups of population including susceptible, expose, infected and recovered are developed. The exercise of studying the model performance against data abundance suggests the modelling results remain highly robust in data sparse setting that is particularly important, for example, in Low-or Middle-Income Country (LMIC).

## Materials and methods

### Data sources

We used publicly available epidemiological data to conduct the analysis, as described below.

#### ONS estimated incidence

Office for National Statistics (ONS) launched Coronavirus (COVID-19) Infection Survey in England on 26 April 2020 to estimate how many people across England, Wales, Northern Ireland and Scotland would have tested positive for COVID-19 infection, regardless of whether they report experiencing symptoms that is one of the primary goals of the survey. The survey is based on a random sample of households to provide a nationally representative survey. Everyone aged 2 years and over in each household sample was asked to take a nose and throat swab for SARS-CoV-2 using reverse transcriptase polymerase chain reaction (RT-PCR). Every participant is swabbed once. they are then invited to have repeat tests every week for another four weeks and then monthly. More descriptions about the survey design can be found [23]. Using Bayesian multilevel generalised additive regression model to model the swab test result (positive or negative) as a function of age, sex, time, and region, the study estimated community prevalence of SARS-CoV-2 in England since April 2020 [10]. Combine the estimates of community prevalence and estimates of duration of PCR testing positivity, the survey modelling team also published the estimates of daily incidence based on a deconvolution model [23].

To conduct the comparison of estimates of incidence between our model and ONS survey, we retrieved the SARS-CoV-2 daily incidence in England in 2020 from the Office for National Statistics (ONS) [11] on March 17, 2023 as shown in Figure 1.

#### Model estimated exposure

Cumulative exposure to SARS-CoV-2 in seven regions of England estimated by the model that we published before were obtained from [1]. Here, we firstly transformed and aggregated the cumulative exposure by region of England to daily incidence in England using Equation (1) and Equation (2).

#### 7-day average of reported COVID-19 cases in England

7-day average of reported COVID-19 daily cases in England in 2020 were retrieved from the UK government’s official COVID-9 online dashboard [12] on March 17, 2023 as shown in Figure 3.

## Method

We firstly calculated the incidence in England estimated by exposure model [1] by computing the difference of cumulative exposure in two successive days and adding together to the whole England as shown in Figure 1 and Figure 3:

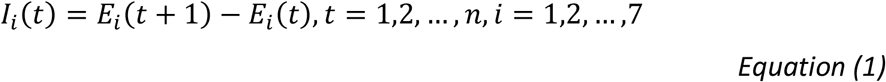

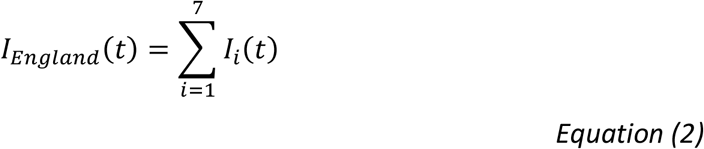

Here, *E*_*i*_(*t*) is the daily exposure at region *i* estimated by exposure model [1], *n* is the total number of days from 1 January 2020 to 7 November 2020, *i* = 1, … 7 represents London, Southwest, Southeast, Northeast, Northwest, East, Midland. *l*_*England*_(*t*) represents the daily incidence of England.

The 7-day average model predicted incidence can be calculated by

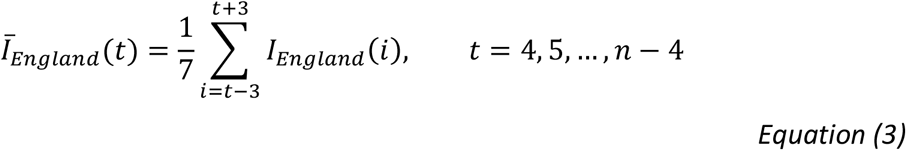

Here, *t* = 4 refers to the fourth day of 2020, *n* is the end date of the comparison exercise, 7 November 2020.

The estimated reporting ratio as shown in Figure 3 was calculated by

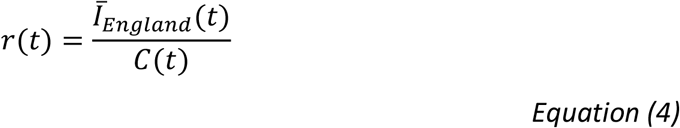

Here, *C* is the 7-day average reported cases in England from the UK government’s official COVID-9 online dashboard [12].

While testing the relationship between model performance and data abundance in Figure 2, we firstly obtained all the data and codes from paper [1] and rerun the model by adding the seroprevalence measurements one by one into the model.

## Data Availability

All data produced in the present work are contained in the manuscript

## Acknowledgments

The authors received no financial support for the research.

## Author contributions

L.J.W., S.C, J.A.F, and K.A.L conceived and designed the study. S.C. cleaned the data, S.C. and L.J.W. developed the methodology and conducted the formal analysis. S.C. and L.J.W. wrote the original manuscript. All authors reviewed and provided analytical input and approved the manuscript.

## Data and materials availability

All codes and materials used in the analyses can be accessed at: https://github.com/SiyuChenOxf/Exposure_ONS-modelling. All parameter estimates and figures presented can be reproduced using the code provided. This work is licensed under a Creative Commons Attribution 4.0 International (CC BY 4.0) license, which permits unrestricted use, distribution, and reproduction in any medium, provided the original work is properly cited.

## References

1. Chen, S., et al., Levels of SARS-CoV-2 population exposure are considerably higher than suggested by seroprevalence surveys. PLOS Computational Biology, 2021. 17(9): p. e1009436.

2. Aburto, J.M., et al., Quantifying impacts of the COVID-19 pandemic through life-expectancy losses: a population-level study of 29 countries. International journal of epidemiology, 2022. 51(1): p. 63–74.

3. Ozili, P.K. and T. Arun, Spillover of COVID-19: impact on the Global Economy, in Managing Inflation and Supply Chain Disruptions in the Global Economy. 2023, IGI Global. p. 41–61.

4. Metcalf, C.J.E., D.H. Morris, and S.W. Park, Mathematical models to guide pandemic response. Science, 2020. 369(6502): p. 368–369.

5. Aguas, R., et al., Modelling the COVID-19 pandemic in context: an international participatory approach. BMJ global health, 2020. 5(12): p. e003126.

6. Pagel, C. and C.A. Yates, Role of mathematical modelling in future pandemic response policy. bmj, 2022. 378.

7. Bollyky, T.J., et al., Pandemic preparedness and COVID-19: an exploratory analysis of infection and fatality rates, and contextual factors associated with preparedness in 177 countries, from Jan 1, 2020, to Sept 30, 2021. The Lancet, 2022. 399(10334): p. 1489–1512.

8. Kennedy, B., et al., App-based COVID-19 syndromic surveillance and prediction of hospital admissions in COVID Symptom Study Sweden. Nature Communications, 2022. 13(1): p. 2110.

9. Desjardins, M.R., Syndromic surveillance of COVID-19 using crowdsourced data. The Lancet Regional Health–Western Pacific, 2020. 4.

10. Pouwels, K.B., et al., Community prevalence of SARS-CoV-2 in England from April to November, 2020: results from the ONS Coronavirus Infection Survey. The Lancet Public Health, 2021. 6(1): p. e30–e38.

11. Office of National Statisics Coronavirus (COVID-19) Infection Survey UK, 2023 https://www.ons.gov.uk/peoplepopulationandcommunity/healthandsocialcare/conditionsanddiseases/bulletins/coronaviruscovid19infectionsurveypilot/latest#strengths-and-limitations. 2023.

12. GOV.UK. Coronavirus (COVID-19) in the UK 2023; Available from: https://coronavirus.data.gov.uk/details/deaths?areaType=nation&areaName=England.

13. Clapham, H., et al., Seroepidemiologic study designs for determining SARS-COV-2 transmission and immunity. Emerging Infectious Diseases, 2020. 26(9): p. 1978.

14. Long, Q.-x., et al., Antibody responses to SARS-CoV-2 in COVID-19 patients: the perspective application of serological tests in clinical practice. MedRxiv, 2020: p. 2020.03.18.20038018.

15. Ibarrondo, F.J., et al., Rapid decay of anti–SARS-CoV-2 antibodies in persons with mild Covid-19. New England Journal of Medicine, 2020. 383(11): p. 1085–1087.

16. Wei, J., et al., Anti-spike antibody response to natural SARS-CoV-2 infection in the general population. Nature Communications, 2021. 12(1): p. 6250.

17. Böger, B., et al., Systematic review with meta-analysis of the accuracy of diagnostic tests for COVID-19. American journal of infection control, 2021. 49(1): p. 21–29.

18. Van Elslande, J., et al., Estimated half-life of SARS-CoV-2 anti-spike antibodies more than double the half-life of anti-nucleocapsid antibodies in healthcare workers. Clinical Infectious Diseases, 2021. 73(12): p. 2366–2368.

19. Knock, E.S., et al., Key epidemiological drivers and impact of interventions in the 2020 SARS-CoV-2 epidemic in England. Science Translational Medicine, 2021. 13(602): p. eabg4262.

20. Russell, T.W., et al., Reconstructing the early global dynamics of under-ascertained COVID-19 cases and infections. BMC medicine, 2020. 18(1): p. 1–9.

21. Chen, S., et al., Estimating the effectiveness of shielding during pregnancy against SARS-CoV-2 in New York City during the first year of the COVID-19 pandemic. Viruses, 2022. 14(11): p. 2408.

22. Bubar, K.M., et al., Model-informed COVID-19 vaccine prioritization strategies by age and serostatus. Science, 2021. 371(6532): p. 916–921.

23. Coronavirus (COVID-19) Infection Survey technical article: Cumulative incidence of the number of people who have tested positive for COVID-19, UK: 22 April 2022. 2020.

